# Factors affecting the choice of a future medical specialty of the junior doctors in a third world country

**DOI:** 10.1101/2021.10.04.21264501

**Authors:** Ifrah J. Malik, Ahsan Tameez-ud-din, Asim Tameez Ud Din, Farooq Mohyud Din

## Abstract

**Background and Objective:** Pakistan is facing a major brain drain and as long as there is not a better understanding of the needs and desires of the junior doctors, this exodus towards the greener pastures shall continue. This study is an effort to recognize the factors which influence the choice of a future specialty of young Pakistani doctors in order to help identify the areas which need improvement.

**Study design:** This descriptive cross-sectional study was conducted from 12th May, 2021 to 2nd August, 2021. Young doctors who had completed their one-year internship in hospitals of Punjab were invited to fill the questionnaire via social media platforms. Data were entered and analyzed using the Statistical Package for Social Sciences (SPSS) version 23.0. Chi-square tests were applied for qualitative variables. A p-value of less than 0.05 was considered significant.

**Results:** Out of a total 105 participants, 60 (57.1%) were females. More females as compared to males decided the future specialty based on work-life balance (20/27, p= 0.039). Internal medicine and general surgery were the most sought-after fields both before and after the internship. Forty-three (41%) participants wished to change their choice of specialty after their house job experiences. Fifty-eight (55.2%) participants considered the future prospects of the field while deciding their specialty while sixty-three (60%) reported the attitude of the senior doctors as an important deciding factor. The Covid-19 pandemic had an effect on 12 (11.4%) participants’ decision regarding their choice of future specialty.

**Conclusion:** Many elements such as internship play a pivotal role in helping the young doctors to narrow their choices. It is important to understand the factors considered by young doctors during their choice of a medical specialty to ensure that a significant proportion of medical work force does not slip through the cracks in our health infrastructure.

## 1. INTRODUCTION

British colonizers are largely credited with the introduction of Western medicine in the Indian subcontinent through the arrival of medical officers onboard the first fleets of East India Company ships [1]. Over the next couple of centuries, the health infrastructure in the subcontinent grew substantially but this growth was not uniform which is evident by the fact that that at the time of independence from the British *raj*, Pakistan (a part of Indian subcontinent before 1947) had just two medical colleges and the services they provided were grossly insufficient to cater to the burgeoning health needs of a new country [2].

College of Physicians and Surgeons of Pakistan (CPSP) was established in 1962 in order to provide a structured approach to Pakistani physicians’ training. Unsurprisingly, the training infrastructure was largely fashioned according to the British health care framework. Over the decades, the Western medical world has undergone many reforms but there have been no major improvements in the Pakistani health sector and the health professionals (including resident doctors in different medical specialties) face a lot of challenges due to a lack of resources and poor utilization of primary health care system [2-4].

The choice of specialty for post graduate medical training is arguably the most important decision of a doctor’s professional life and there are many factors which affect this selection process [5,6]. In a resource-limited country like Pakistan, the choice of specialty may be influenced by additional sociocultural considerations like family pressure and lack of training facilities. The role of house job (a one-year mandatory hospital internship before medical graduates can apply for residency) in the growth of health professionals has often been discussed in health conferences and international publications, but there is a scarcity of literature describing its part in a young physician’s selection of medical field [7]. The policy makers and senior health executives need to have a better understanding of the factors that influence the choice of medical specialties in order to help the young doctors navigate this turbulent and uncertain period of their lives in a stress-free manner.

Pakistan is facing a major brain drain as many medical graduates are leaving the country for better job opportunities abroad [8]. The arduous path to acquiring a residency spot in their favorite specialty, difficult working conditions and disappointing prospects of the medical fields in the country are some common complaints and as long as there is not a better understanding of the needs and desires of the young doctors, this exodus towards the greener pastures shall continue. This study is an effort to recognize the factors which influence the choice of a future specialty of young Pakistani doctors in order to help provide a framework for the policy makers to identify the areas which need improvement and make impactful decisions in this regard. Our study also uniquely describes the impact of the mandatory hospital internship over the decision-making process and explores whether the pandemic has had any influence over the selection of a specialty. Figure 1 shows a simplified comparison of the likely professional journey of a medical graduate in Pakistan, United Kingdom and United States of America.

**Figure 1:**
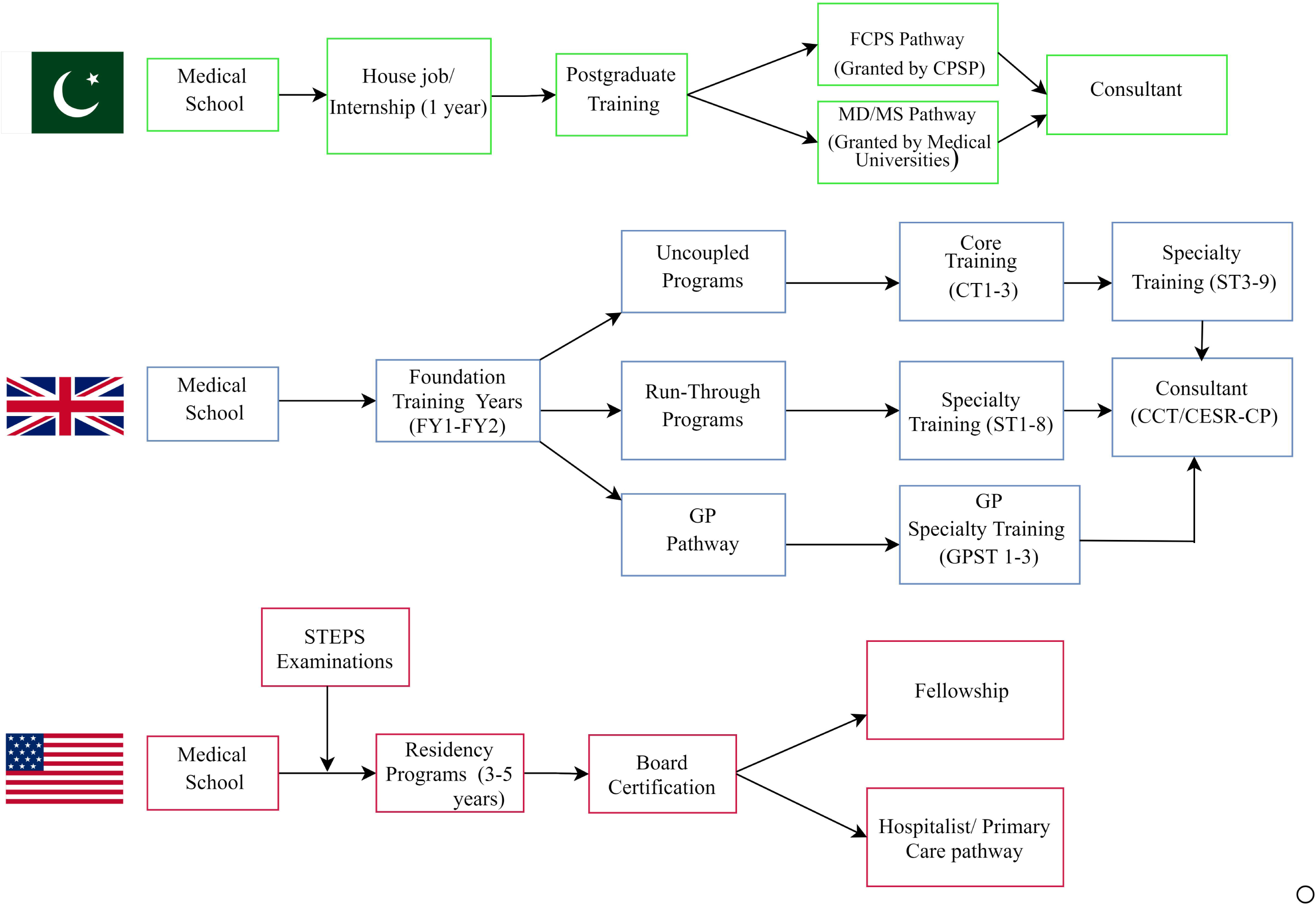
shows an outline of a simplified pathway of a medical graduate to becoming a consultant in Pakistan, United Kingdom (UK) and United States of America (USA) Abbreviations: FCPS- Fellow of College of Physicians and Surgeons Pakistan, CPSP- College of Physicians and Surgeons Pakistan, MD-Doctor of Medicine, MS- Master of Surgery, CCT- Certificate of Completion of Training, CESR- Certificate of Eligibility for Specialist Registrar, CP-Combined Program

## 2. MATERIALS AND METHODS

### 2.1 Scope of study

This descriptive cross-sectional study was conducted from 12^th^ May,2021 to 2^nd^ August,2021. Ethical approval was obtained from the ethical review board (ERB) of Rawalpindi Medical University. Young doctors who had completed their internship in private and public sector tertiary care hospitals of the Punjab province of Pakistan were invited to fill the questionnaire in person or via social media platforms.

### 2.2 Sampling technique

The data was collected by convenience sampling technique. The doctors who agreed to take part in the study were provided with a link to the questionnaire. Those who had completed their house job within the last 6 months were included in the study while the doctors who had started their residency were excluded.

### 2.3 Questionnaire design

The questionnaire consisted of two parts. The first part comprised of questions regarding the demographic details of the doctors while the second part had queries concerning the different factors impacting their decision of a future specialty. Towards the end of the questionnaire, a couple of optional questions regarding the impact of the COVID-19 pandemic were asked.

### 2.4 Data analysis

Data were entered and analyzed using the Statistical Package for Social Sciences (SPSS) version 23.0 (IBM Corp, Armonk, US). The results were reported as frequencies, percentages, and figures. Chi-square tests were applied for qualitative variables. A p-value of less than 0.05 was considered significant.

## 3. RESULTS

Out of a total 105 participants, 60 (57.1%) were females while the rest [45 (42.9%)] were males. Most of them (83, 79%) had graduated from government institutes, 18 (17.1%) were graduates of private institutes whereas four (3.8%) were foreign medical graduates. Eighty-two (78.1%) participants had done their internship from the hospital affiliated with their parent institute of graduation while 23 (21.9%) had chosen a hospital not affiliated with their parent institute for their internship.

Some of the factors affecting their choice of specialty are shown in **Table 1**. More females as compared to males decided the future specialty based on work-life balance (20/27, p= 0.039). Most of the participants who thought that exposure to clinical rotations impacted their choice of future specialties were also affected by the attitude and professionalism of their senior doctors (16/20, P= 0.042).

**Table 1:** Factors considered by junior doctors while deciding a medical field.

| **Do you consider the following factors while deciding your future medical field?** | **YES**<br>**n (%)**<br>**(N= 105)** | **NO**<br>**n (%)**<br>**(N= 105)** |
| --- | --- | --- |
| Scope of the field | 43 (41) | 62 (59) |
| Working environment | 20 (19) | 85 (81) |
| Interest of friends/family | 20 (19) | 85 (81) |
| Work/life balance | 27 (25.7) | 78 (74.3) |
| Diversity/challenging nature of job | 34 (32.4) | 71 (67.6) |
| Exposure during ward rotations in 3rd-5th years | 20 (19) | 85 (81) |
| Good pay package | 9 (8.6) | 96 (91.4) |

Thirty-seven (35.2%) participants reported that their choice of specialty was influenced by factors such as family pressure. Fifty-eight (55.2%) participants said that the future prospects of specialty in their country affected their choice. Seventy-seven (73.3%) participants did their rotation (during house job) in the specialty they wanted to pursue while 28 (26.7%) did not. Sixty-eight (64.8%) had a specific center in mind where they wanted to complete their residency and 65 (61.9%) considered the available facilities while selecting their center for residency. Sixty-three (60%) participants reported that the attitude of their seniors and 35 (33.3%) reported that the attitude of paramedic staff affected their decision about selecting their specialty of choice. Ninety-eight (93.3%) held the opinion that different personality types are suited for different medical specialties. Twenty-five (23.8%) participants reported that their decision of choosing a particular specialty was influenced by a family member already working in a specific field. This is shown in **Table 2**.

**Table 2:** Questions related to the impact of different circumstances and factors while deciding a medical specialty.

| Questions | Response | Frequency (%) |
| --- | --- | --- |
| Did the future prospects of this field in your country made you consider it? | Yes | 58 (55.2%) |
|  | No | 47 (44.8%) |
| Did the attitude/professionalism of your seniors have an impact on your choice of speciality? | Yes | 63 (60%) |
|  | No | 42 (40%) |
| Did your experience with supporting staff (nurses, ward boys etc) impact your decision in any way? | Yes | 35 (33.3%) |
|  | No | 70 (66.7%) |
| Did you consider the facilities available as an important decision-making factor in your choice of training centre? | Yes | 65 (61.9%) |
|  | No | 40 (38.1%) |
| Do you think different personality types are suited for different medical specialities/subspecialties? | Yes | 98 (93.3%) |
|  | No | 7 (6.7%) |
| Did the presence of your friend/family member in a specific medical speciality influence your decision in any way? | Yes | 25 (23.8%) |
|  | No | 80 (76.2%) |

### 3.1 Effect of House Job (1-year mandatory internship)

Forty-one (39%) participants reported that they had chosen internal medicine as their future specialty before their house job while 29 (27.6%) said that they had an interest in surgery before the internship. Ten (9.5%) internees had chosen Radiology, Ophthalmology, Anesthesia and Dermatology (R.O.A.D. specialties) as their future specialty of choice. Sixty-two (59%) participants wanted to pursue the same medical specialty after completing their house job whereas 43 (41%) wished to change it after house job. The breakdown of specialties of choice before and after house job is shown in **Table 3**.

**Table 3:** comparison of the choice of field before and after House job.

| CHOICE BEFORE HOUSE JOB |  | CHOICE AFTER HOUSE JOB |  |
| --- | --- | --- | --- |
| Speciality | Frequency (%) | Speciality | Frequency (%) |
| Internal Medicine | 41 (39) | Internal Medicine | 33 (31.4) |
| Emergency Medicine | 2 (1.9) | Emergency Medicine | 3 (2.9) |
| Rheumatology | 1 (1) | Rheumatology | 1 (1) |
| Psychiatry | 4 (3.8) | Psychiatry | 1 (1) |
| Paediatrics | 2 (1.9) | Paediatrics | 3 (2.9) |
| General Surgery | 29 (27.6) | General Surgery | 29 (27.6) |
| Otorhinolaryngology | 1 (1) | Otorhinolaryngology | 3 (2.9) |
| Neurosurgery/Neurology | 3 (2.9) | Neurosurgery/Neurology | 2 (1.9) |
| Cardiology/Cardiothoracic surgery | 3 (2.9) | Cardiology/Cardiothoracic surgery | 4 (3.8) |
| Gynaecology | 4 (3.8) | Gynaecology | 3 (2.9) |
| Orthopaedics | 2 (1.9) | Orthopaedics | 2 (1.9) |
| R.O.A.D. | 10 (9.5) | R.O.A.D. | 17 (16.2) |
| None | 3 (2.9) | None | 1 (1) |

Most of the participants (101, 96.2%) were of the opinion that house job is mandatory for growth of medical professionals, while only 4 (3.8%) said otherwise. The role of this internship period in the professional growth is further elucidated in **Table 4**.

**Table 4:**
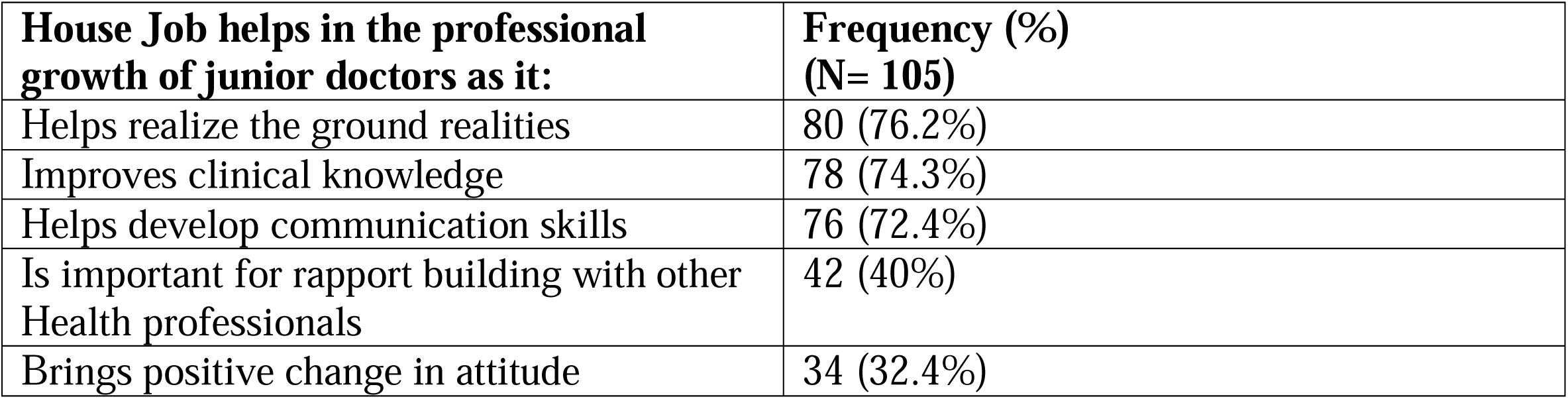
Role of house job in the professional growth of junior doctors.

| **House Job helps in the professional growth of junior doctors as it:** | **Frequency (%)**<br>**(N= 105)** |
| --- | --- |
| Helps realize the ground realities | 80 (76.2%) |
| Improves clinical knowledge | 78 (74.3%) |
| Helps develop communication skills | 76 (72.4%) |
| Is important for rapport building with other Health professionals | 42 (40%) |
| Brings positive change in attitude | 34 (32.4%) |

### 3.2 Effect of the Covid-19 pandemic

Eighty-four (80%) participants had worked in Covid-19 wards during their rotations while 21 (20%) had not. The number of participants who suffered from Covid-19 infection during their house job was 59 (56.2%) whereas 46 (43.8%) did not contract the virus. The pandemic had an effect on 12 (11.4%) participants’ decision regarding their choice of specialty and only 36 (34.3%) participant wanted to pursue a medical field with high risk of exposure to infectious pathogens.

## 4. DISCUSSION

Post graduate training is an essential rite of passage during the professional lives of most doctors and the learning experiences during this stage are widely considered to shape their future teaching and clinical practices. The resident doctors make up a large proportion of the essential work force of the hospitals and they are the front-line warriors in any health emergency or natural disaster [9]. The Covid-19 pandemic has further cemented the importance of post graduate medical trainees and the positive role played by these doctors has been recognized by people from all over the world [10,11].

In spite of the undeniable significance of this phase of a doctor’s life, the health authorities in our country have not paid much attention towards the needs and desires of the junior doctors which is resulting in an exodus of brilliant physicians and surgeons from the country [8,12]. The choice of a post graduate medical specialty is a crucial decision in the young physicians’ lives and the authorities ought to recognize the factors that influence this choice as well as the expectations of the aspiring specialists from their place of residency.

A majority of the participants in our study were females (57.1%) which is unsurprising as the number of female medical graduates in the country far outnumbers their male counterparts [13]. A significantly larger proportion of female participants considered work/life balance as an important deciding factor in the choice of their future specialty (p = 0.039). The health authorities have long raised concerns regarding the low percentage of the female medical graduates later contributing towards the overall work force of the hospitals due to social issues such as arranged marriage and cultural taboos related to the working women [14,15]. A change in the overall patriarchal mindset of the society and the deeply entrenched gender roles is the need of hour in order to stop this valuable proportion of the medical work force from going to waste.

Most of the junior doctors had considered internal medicine as their future specialty before house job followed by general surgery (39% and 27.6% respectively) and these fields were also the most popular choice after their one-year mandatory internship (31.4% and 27.6% respectively). This was similar to the findings of other studies from the country [6,16]. However, more than one third of the doctors approached (41%) said that their choice of future specialty had changed after their house job experiences which indicated the crucial role this period plays in the selection of a future medical field. A whopping majority of the participants (96.2%) were off the view that house job has a positive impact on the professional growth of junior doctors. Almost 76% of the participants recognized the role played by this period in helping the doctors to realize the ground realities, while 74.3% and 72.4% agreed that it helps in improving the clinical knowledge and developing communication skills of doctors respectively. No one can deny the importance of communication skills in the field of medicine and in countries with poor quality of school education, such a phase of practical exposure becomes all the more important for the growth of individuals including doctors [17-19]. In short, although the importance of mandatory internship before residency (known as foundation years in UK and house job in Pakistan) in the choice of future specialty and the growth of health professionals cannot be overstated, further work needs to be done in order to maximize the learning possibilities this phase has to offer, in order to help nurture a generation of safe, confident and skilled doctors [20-22].

Sixty percent of the participants in our study considered that the attitude and professionalism of the senior doctors and faculty members was important in their choice of a certain medical specialty. A significant proportion of the doctors, who were of the opinion that the exposure of clinical rotations impacted their choice of future specialty, were also influenced in their decision by the attitude and professionalism of their senior doctors (p = 0.042). Interestingly, a vast majority (91.4%) did not consider the salary package to be an important deciding factor in their selection. More than 64 percent of the internees had a specific center in mind where they want to complete their residency in future and the available training facilities were deemed important in the choice of the center (61.9%). Nagler et al. reported that the relationship between the faculty and current residents in a specialty as well as the quality of training were very important factors considered by applicants during their choice of a training program while salary was not a major consideration [23]. Similar observations have been reported by other studies which underscore the importance of a positive interaction between senior doctors and their juniors during the medical school ward rotations and later the internship period [24]. Moreover, such findings emphasize the importance of uniform and comprehensive availability of training facilities in order to optimize the opportunities available to the junior doctors during their search for a suitable training program.

Specialty choice in medicine has often been associated with the type of personality of the individuals and a vast majority of the junior doctors in our study (93.3%) agreed with this assumption [25]. Covid-19 pandemic is an unparalleled health disaster of the modern times and has caused seismic shifts in the delivery of health-care facilities [26,27]. Junior doctors have been at the forefront in the fight against this novel disease and it is important to assess the impact of the pandemic over their future decisions. Eighty percent of our study participants had performed duties in Covid-19 wards during their house job. A majority of the participants (56.2%) reported that they had suffered from the disease during their hospital duties which underlines the risk faced by the frontline health workers during this pandemic and it has been corroborated by many international studies [28,29]. Despite the profound effects of Covid-19 on the delivery of global health-care services, only 11.4% of the junior doctors in our study agreed that the pandemic had an effect on their choice of a future specialty. The long-term impact of this disease remains to be seen and there is a deficiency of literature regarding this topic [30]. Further studies will be required in the future in order to map the effect of such a global disaster over the choices of the future generation of doctors.

### Limitation

The main limitation of our study was that the data from the junior doctors working in other provinces of Pakistan could not be gathered. Although, Punjab is the largest province (by population) of Pakistan, the data from other provinces may have revealed some significant differences owing to the certain geographic and cultural peculiarities specific to every province.

## 5. CONCLUSION

This study provides insight into the budding minds of junior doctors of Pakistan and indicates the factors they consider important while deciding a medical specialty. Most medical graduates have a pervasive sense of uncertainty regarding their future career in medicine and internship has a pivotal role to play in helping them narrow their choices. Empathetic and kind supervision and a stress-free environment can make it easier to pick a probable path that suits their temperament. There is also a need to make the medical profession more gender inclusive in order to help ease most of the female doctors into the specialties of their choice. It is the need of the hour to understand the factors considered by young doctors during their choice of a medical specialty to ensure that a significant proportion of medical work force does not slip through the cracks in our health infrastructure and everyone can play their part in the improvement of the health sector of this country as well as contributing towards global health efforts.

## Data Availability

Relevant data is included in the manuscript. Additional data may be provided upon reasonable request.

## c. Acknowledgements

None.

## REFERENCES

1. Anshu AS. Evolution of medical education in India: The impact of colonialism. Journal of postgraduate medicine. 2016 Oct;62(4):255. DOI: 10.4103/0022-3859.191011

2. Pulsepakistan.com. 2021. Postgraduate Medical Education in Pakistan (1947 to 2012). [online] Available at: <http://www.pulsepakistan.com/index.php/main-news-nov-1-12/104-postgraduate-medical-education-in-pakistan-1947-to-2012> [Accessed 30 July 2021].

3. Biggs JS. Postgraduate medical training in Pakistan: observations and recommendations. Journal of the College of Physicians and Surgeons Pakistan.. 2008;18(1):58–63. URL:https://www.jcpsp.pk/archive/2008/Jan2008/17.pdf [Accessed30 July 2021].

4. Khalid F, Abbasi AN. Challenges faced by Pakistani healthcare system: Clinician’s perspective. Journal of the College of Physicians and Surgeons Pakistan. 2018;28(12):899–901 URL: https://www.jcpsp.pk/archive/2018/Dec2018/01.pdf

5. Nagler A, Andolsek K, Schlueter J, Weinerth J. To match or not: factors influencing resident choice of graduate medical education program. Journal of graduate medical education. 2012;4(2):159-64. DOI: https://doi.org/10.4300/JGME-D-11-00109.1

6. Aslam M, Ali A, Taj T, Badar N, Mirza W, Ammar A, Muzaffar S, Kauten JR. Specialty choices of medical students and house officers in Karachi, Pakistan. EMHJ-Eastern Mediterranean Health Journal.2011;17(1):74–79. URL: https://applications.emro.who.int/emhj/V17/01/17_1_2011_0074_0079.pdf

7. Yousuf S, Siddiqui N. Learning procedures during house job. Journal of the Pakistan Medical Association. 2008;58(12):698. URL:https://pubmed.ncbi.nlm.nih.gov/19157326/

8. Imran N, Azeem Z, Haider II, Amjad N, Bhatti MR. Brain drain: post graduation migration intentions and the influencing factors among medical graduates from Lahore, Pakistan. BMC research notes. 2011;4(1):1–5. DOI: https://doi.org/10.1186/1756-0500-4-417

9. van der Leeuw RM, Lombarts KM, Arah OA, Heineman MJ. A systematic review of the effects of residency training on patient outcomes. BMC medicine. 2012;10(1):1–1. DOI: https://doi.org/10.1186/1741-7015-10-65

10. The vital role residents play in health care during pandemic [Internet]. American Medical Association. 2021 [cited 3 September 2021]. Available from: https://www.ama-assn.org/residents-students/residency/vital-role-residents-play-health-care-during-pandemic

11. Baptista FV, Aguiar MR, Moreira JA, Sousa FC, Plenns GC, Simao RR, Ruffini VM, Lin CA, Nunes MD. Contributions of residents from multiple specializations in managing the COVID-19 pandemic in the largest public hospital Brazil. Clinics. 2020;75. DOI: https://doi.org/10.6061/clinics/2020/e2229

12. Hossain N, Shah N, Shah T, Lateef SB. Physicians’ migration: perceptions of Pakistani medical students. J Coll Physicians Surg Pak. 2016;26(8):696-701.URL: https://pubmed.ncbi.nlm.nih.gov/27539766/

13. Pakistan’s medical schools: where the women rule [Internet]. DAWN.COM. 2013 [Accessed: 4 September 2021]. Available from: https://www.dawn.com/news/803667/pakistans-medicalschools-where-the-women-rule

14. Moazam F, Shekhani S. Why women go to medical college but fail to practise medicine: perspectives from the Islamic Republic of Pakistan. Medical education. 2018;52(7):705-15.DOI: 10.1111/medu.13545

15. Masood A. Influence of Marriage on Women’s Participation in Medicine: The Case of Doctor Brides of Pakistan. Sex Roles. 2019;80.DOI:https://doi.org/10.1007/s11199-018-0909-5

16. Ali A, Rasheed A, Zaidi SM, Alsaani SM, Naim H, Hamid H, Farrukh S. Recent Trend in Specialty Choices of Medical Students and House Officers from Public Sector Medical Universities, Karachi. JPMA. 2019;69(4):489–494. URL: https://pubmed.ncbi.nlm.nih.gov/31000850/

17. Amanat R, Yasmin M, Sohail A, Amanat M. Pakistani Medical Students’ Attitudes towards Communication Skills Learning: A Correlation of Demographic and Education-Related Characteristics. Open Journal of Social Sciences. 2016;4(01):67.DOI: 10.4236/jss.2016.41009

18. Ferreira-Padilla G, Ferrández-Antón T, Baleriola-Júlvez J, Braš M, Đorđević V. Communication skills in medicine: where do we come from and where are we going?. Croatian medical journal. 2015;56(3):311.DOI: 10.3325/cmj.2015.56.311

19. Ghazi SR, Ali R, Khan MS, Hussain S, Fatima ZT. Causes of the decline of education in Pakistan and its remedies. Journal of College Teaching & Learning (TLC). 2010;7(8).DOI: https://doi.org/10.19030/tlc.v7i8.139

20. Collins H, Eley C, Kohler G, Morgan H. Foundation rotations in medical training: is it love at first sight?. Postgraduate Medical Journal. 2021 May 26. DOI: 10.1136/postgradmedj-2021-140198

21. Dean BJ, Duggleby PM. Foundation doctors’ experience of their training: a questionnaire study. JRSM short reports. 2013;4(1):1–7.DOI: 10.1258/shorts.2012.012095

22. Ahmad A, Su HS, Tayyab M, Gardezi JR. Perceptions of doctors on being treated by a doctor just completing the house job. Journal of the College of Physicians and Surgeons--pakistan: JCPSP. 2014;24(12):908-13. URL: https://pubmed.ncbi.nlm.nih.gov/25523726/

23. Nagler A, Andolsek K, Schlueter J, Weinerth J. To match or not: factors influencing resident choice of graduate medical education program. Journal of graduate medical education. 2012;4(2):159–64.DOI: 10.4300/JGME-D-11-00109.1

24. Sinno S, Mehta K, Squitieri L, Ranganathan K, Koeckert MS, Patel A, Saadeh PB, Thanik V. Residency characteristics that matter most to plastic surgery applicants: a multi-institutional analysis and review of the literature. Annals of plastic surgery. 2015;74(6):713–7.DOI: 10.1097/SAP.0000000000000511.

25. Al-Ansari SS, Khafagy MA. Factors affecting the choice of health specialty by medical graduates. Journal of family & community medicine. 2006;13(3):119. URL: https://www.ncbi.nlm.nih.gov/pmc/articles/PMC3410059/

26. Arabi YM, Azoulay E, Al-Dorzi HM, Phua J, Salluh J, Binnie A, Hodgson C, Angus DC, Cecconi M, Du B, Fowler R. How the COVID-19 pandemic will change the future of critical care. Intensive care medicine. 2021:282–291. DOI:https://doi.org/10.1007/s00134-021-06352-y

27. Yager PH, Whalen KA, Cummings BM. Repurposing a pediatric ICU for adults. New England Journal of Medicine. 2020;382(22):e80. DOI:

28. Nguyen LH, Drew DA, Graham MS, Joshi AD, Guo CG, Ma W, Mehta RS, Warner ET, Sikavi DR, Lo CH, Kwon S. Risk of COVID-19 among front-line health-care workers and the general community: a prospective cohort study. The Lancet Public Health. 2020;5(9):e475-83. DOI: https://doi.org/10.1016/S2468-2667(20)30164-X

29. Atnafie SA, Anteneh DA, Yimenu DK, Kifle ZD. Assessment of exposure risks to COVID-19 among frontline health care workers in Amhara Region, Ethiopia: A cross-sectional survey. Plos one. 2021;16(4):e0251000. DOI: https://doi.org/10.1371/journal.pone.0251000

30. Deng J, Que J, Wu S, Zhang Y, Liu J, Chen S, Wu Y, Gong Y, Sun S, Yuan K, Bao Y. Effects of COVID-19 on career and specialty choices among Chinese medical students. Medical Education Online. 2021;26(1):1913785. DOI: https://doi.org/10.1080/10872981.2021.1913785

